# Health Information Seeking Experiences Among People with Disabilities: Results from the HINTS 2024

**DOI:** 10.1101/2025.10.14.25338014

**Authors:** Maria Velat, Tyler G. James

## Abstract

**Background:** People with disabilities (PWDs) face disparities in the healthcare system that lead to poorer health outcomes. A lack of health information and accessible communication with healthcare professionals is linked to these health inequities. PWDs report lower health literacy, technology use, and different access needs that limit effective health-related communication. Due to the broad spectrum of disabilities, the barriers PWDs face with accessing health information vary greatly.

**Methods:** We conducted an existing data analysis using the nationally representative Health Information National Trends Survey (HINTS) conducted in 2024. We estimated adjusted odds ratios (aORs) using logistic regression for five different cancer-related health information outcomes among US civilian, non-institutionalized adults who included disability status (weighted n=250,488,318).

**Results:** Primary findings indicated that PWDs—especially those with multiple disabilities, chronic pain, or deafness—had disparities in their health information seeking experiences compared to people without disabilities. People with multiple disabilities had higher odds of reporting frustration and difficulty understanding health information, as well as not searching for health information in the first place.

**Conclusion:** People with disabilities experience barriers to seeking health information but these barriers differ on the type of disability. This study is novel in the ability to compare different types of disabilities to different health information outcomes, but true disability representation is likely not possible due to inaccessible survey design. The findings in this study highlight the need for accessible health information, surveys, and more interventions that include PWDs in public health programming.

## Introduction

One in four people report having a disability in the United States and this number is only expected to grow as the population ages.^1^ People with disabilities (PWDs) face discrimination, especially in healthcare systems, and are twice as likely to develop additional health conditions even as access to healthcare grows worldwide.^2^ Many of these health inequities are connected to a lack of effective communication and access to health information between PWDs and healthcare professionals.^3^ Access to information and effective communication are important determinants of positive outcomes in healthcare settings.^4^ A lack of health information, combined with low health literacy and access to technology among PWDs, leads to preventable health complications for disabled individuals.^5-7^ Current national and global health objectives such as Healthy People 2030 emphasize health information as a public health priority, especially for PWDs.^8^ It is important to use new nationally representative data to assess how health information is affected by disability to further lead policy that aims to decrease health disparities. It is essential that those who are more vulnerable to health complications and who interact with the healthcare system more frequently are able to both access and understand necessary health information. Understanding how PWDs access and interact with health information is an important step in bridging disparities that exist between disabled individuals and their trust in the healthcare system.

The Health Information National Trends Survey (HINTS) is conducted yearly by the National Cancer Institute to assess access to and use of health information related to cancer.^9^ For the first time, the most recent HINTS survey has asked a question that accounts for four different types of disabilities. This allows for differences in health information seeking experiences to be assessed across those with sensory disabilities (i.e., deaf and hard-of-hearing, and blind/low-vision), mobility disabilities, pain-related disabilities, as well as those with multiple disabilities. Experiences in accessing health information differs greatly between different disability types, but all PWDs experience inequities. However, the manifestation of those inequities appears differently depending on the type of disability a person has. For example, those who are deaf or blind/low-vision have different access needs and may need information to be communicated in different modalities.^10-11^ In comparison, people with mobility and pain-related disabilities may not have different information access needs, but they may have lower health and technology literacy that affects their access to and comprehension of health information.^12-13^ Experiences in healthcare also differ depending on the time of onset for a disability, as those who are born with a disability have experienced the health system differently than those with acquired disabilities.

Given the different communication methods between disability types and experiences in the healthcare system, it is expected that access and trust in health information will differ based on type of disability.^4,7^ Prior studies have analyzed literacy and accessibility issues when it comes to accessing health information, but there is a clear gap in the literature analyzing the health information seeking experiences among PWDs.^6-7,9^ Using HINTS questions relating to both disability type and health information, in this study we aim to assess how experiences seeking health information differ between people with and without disabilities, as well as between different disability types. It was hypothesized that individuals with sensory disabilities (i.e., deaf and hard-of-hearing, and blind/low-vision) experience the greatest disparities in their effort and frustration in gathering health information, as well as understanding and trusting its quality.

## Methods

### Data Source

The National Cancer Institute aims to understand health information support needs in the United States in a nationally representative sample through HINTS.^9^ We used data from HINTS 7 (2024) because it included four different options for disabilities for the first time since the survey began in 2003. HINTS 7 was conducted between March 2024-September 2024. The sample of US civilian, non-institutionalized adults was determined by randomly selecting addresses from the US Postal Service file of stratified residential addresses and then selecting the adult in the household whose birthday is next. High-minority and rural/nonmetropolitan areas are routinely oversampled in HINTS.^14^ HINTS 7 had a 27.31% response rate and respondents could choose between a postal and push-to web option. More details on HINTS survey design and procedure is available in the HINTS methodology report.^9^ Of the 7278 respondents, 6609 answered questions about their disability status, which represents 250,488,318 people with weights.

### Measures

#### Outcomes

HINTS questions about seeking health information assessed both the process of seeking information and opinions about the information itself. First respondents were asked if they had ever looked for information about cancer from any source. Respondents who had searched for cancer-related information were asked about four barriers: the search taking a lot of effort, feeling frustrated during the search, being concerned about the quality of information, and difficulty understanding the information found. Response options were: strongly agree, somewhat agree, somewhat disagree, or strongly disagree. These responses were dichotomized to agree (i.e., strongly agree/somewhat agree) or disagree (i.e., strongly disagree/somewhat disagree).

#### Disability status

Unlike other government surveys in the U.S., disability status was not measured using the six American Community Survey items. Instead, participants were asked if the specific condition significantly limited daily activities. We created a mutually exclusive disability variable, including the four original disabilities measured (i.e., deafness, blindness, mobility, and pain), and a fifth ‘multiple’ disability option. Participants who answered “no” to all of the conditions were coded as “none.”

#### Covariates

We used a directed acyclic graph (DAG) to identify confounders that should be adjusted for in the statistical models estimating the total effect of disability status on information barriers; we adjusted for age and ever having cancer, which were identified as confounders. Individual level demographics such as race/ethnicity, income level, and education level are reported; these variables are on the theoretical causal path between disability status and the outcomes and were not adjusted for.

### Data Analysis

All data analyses were conducted in SAS version 9.4. We estimated descriptive statistics using frequencies and percentages for demographic characteristics and outcome variables, in addition to trust of cancer-related information sources. For outcome modeling, we intended to use Mplus to estimate adjusted prevalence ratios using Poisson regression and account for missing data using Full Information Maximum Likelihood (FIML). Due to convergence issues, and as specified in our preregistration, we estimated adjusted odds ratios (aORs) using logistic regression with complete case analysis using PROC SURVEYLOGISTIC in SAS. Each of the five outcomes were modeled using logistic regression, adjusting for ever having cancer and age, with replicate weights. As the four information barriers (i.e., effort, frustration, quality, and understanding) were only asked to those who had ever searched for cancer information, the estimates represent conditional adjusted odds ratios. We adjusted for multiple comparisons using a Holm-Bonferroni adjustment. Sensitivity analyses are described in **Supplemental Appendix 1**.

## Results

Demographic characteristics of the unweighted and weighted sample are available in **Table 1**. The most common disability type was multiple disabilities (unweighted prevalence: 13.61%), followed by chronic pain (unweighted: 8.16%). Among those with multiple disabilities, the majority had a combination of mobility with other disabilities including pain (unweighted: 50.51%) or blindness (unweighted: 8.81%), or mobility with deafness and blindness (unweighted: 8.47%). The sample was predominantly white, with at least some college education. Primary findings are reported in **Table 2**. DHH individuals and those with multiple disabilities had lower aOR of ever searching for cancer-related information (aOR = 0.507; 95% CI: 0.306 to 0.840). Among people who searched for information, people with multiple disabilities had higher odds of reporting frustration (aOR = 1.587; 95% CI: 1.081 to 2.332) and difficulty understanding (aOR = 1.442; 95% CI: 1.041 to 1.998). In addition, people who have chronic pain had higher odds of reporting concerns related to the quality of information (aOR = 1.901; 95% CI: 1.089 to 3.319). There were no significant differences in any of the outcomes for individuals who were BLV or who had mobility disabilities. After multiple comparison adjustment within-outcome family using Holm-Bonferroni adjustment, the only significant association was that DHH people had lower odds of seeking cancer information; this indicates that other apparent disparities may be sensitive to multiple testing.

**Table 1.**
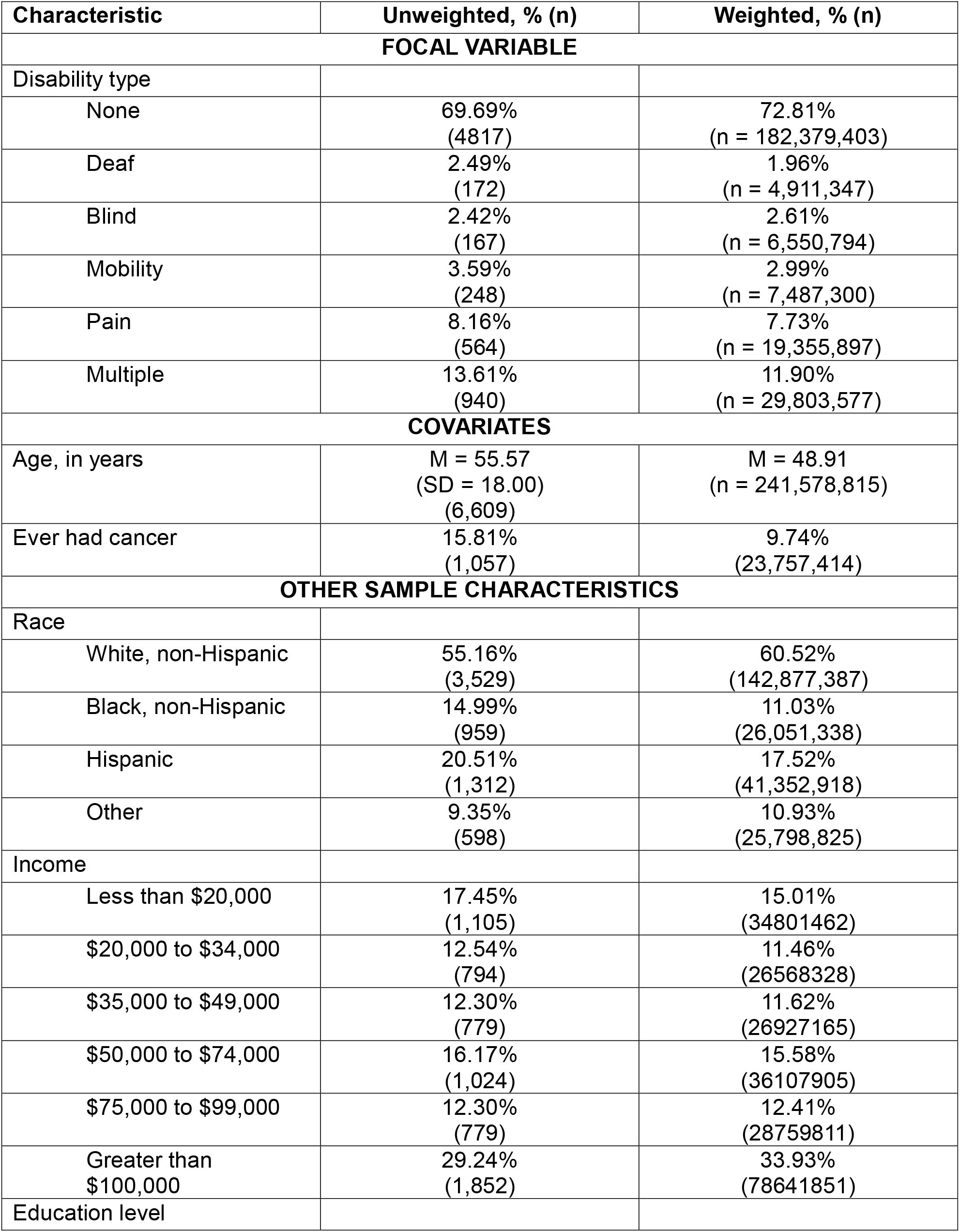
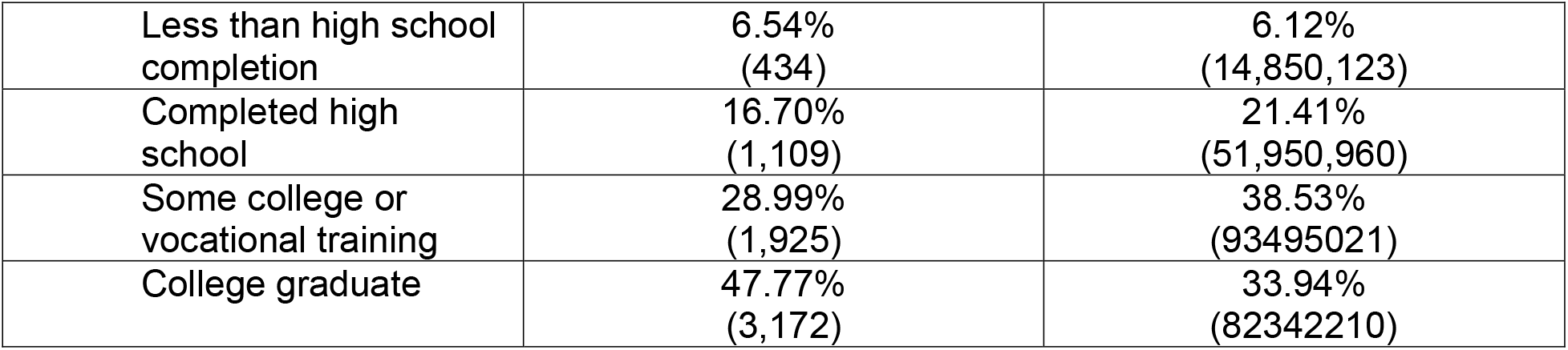
Demographic characteristics.

| Characteristic | Unweighted, % (n) | Weighted, % (n) |
| --- | --- | --- |
| **FOCAL VARIABLE** |  |  |
| Disability type |  |  |
| None | 69.69%<br>(4817) | 72.81%<br>(n = 182,379,403) |
| Deaf | 2.49%<br>(172) | 1.96%<br>(n = 4,911,347) |
| Blind | 2.42%<br>(167) | 2.61%<br>(n = 6,550,794) |
| Mobility | 3.59%<br>(248) | 2.99%<br>(n = 7,487,300) |
| Pain | 8.16%<br>(564) | 7.73%<br>(n = 19,355,897) |
| Multiple | 13.61%<br>(940) | 11.90%<br>(n = 29,803,577) |
| **COVARIATES** |  |  |
| Age, in years | M = 55.57<br>(SD = 18.00)<br>(6,609) | M = 48.91<br>(n = 241,578,815) |
| Ever had cancer | 15.81%<br>(1,057) | 9.74%<br>(23,757,414) |
| **OTHER SAMPLE CHARACTERISTICS** |  |  |
| Race |  |  |
| White, non-Hispanic | 55.16%<br>(3,529) | 60.52%<br>(142,877,387) |
| Black, non-Hispanic | 14.99%<br>(959) | 11.03%<br>(26,051,338) |
| Hispanic | 20.51%<br>(1,312) | 17.52%<br>(41,352,918) |
| Other | 9.35%<br>(598) | 10.93%<br>(25,798,825) |
| Income |  |  |
| Less than \$20,000 | 17.45%<br>(1,105) | 15.01%<br>(34801462) |
| \$20,000 to \$34,000 | 12.54%<br>(794) | 11.46%<br>(26568328) |
| \$35,000 to \$49,000 | 12.30%<br>(779) | 11.62%<br>(26927165) |
| \$50,000 to \$74,000 | 16.17%<br>(1,024) | 15.58%<br>(36107905) |
| \$75,000 to \$99,000 | 12.30%<br>(779) | 12.41%<br>(28759811) |
| Greater than \$100,000 | 29.24%<br>(1,852) | 33.93%<br>(78641851) |
| Education level |  |  |
| Less than high school completion | 6.54%<br>(434) | 6.12%<br>(14,850,123) |
| Completed high school | 16.70%<br>(1,109) | 21.41%<br>(51,950,960) |
| Some college or vocational training | 28.99%<br>(1,925) | 38.53%<br>(93495021) |
| College graduate | 47.77%<br>(3,172) | 33.94%<br>(82342210) |

**Table 2.** Weighted adjusted logistic regression results of health information barriers among people with disabilities in the U.S.

|  | Deaf | Blind | Mobility | Pain | Multiple |
| --- | --- | --- | --- | --- | --- |
| Seeking cancer information<br>(weighted n = 238,636,151) | **0.507**<br>**(0.306 to 0.840)\*** | 0.695<br>(0.330 to 1.464) | 0.870<br>(0.499 to 1.518) | 0.997<br>(0.701 to 1.418) | **0.726**<br>**(0.561 to 0.940)** |
| Effort<br>(weighted n=121,789,507) | 1.004<br>(0.353 to 2.857) | 1.161<br>(0.253 to 5.318) | 1.433<br>(0.655 to 3.134) | 1.198<br>(0.674 to 2.127) | 1.361<br>(0.965 to 1.918) |
| Frustration<br>(weighted n= 121,439,758) | 0.897<br>(0.397 to 2.023) | 0.941<br>(0.234 to 3.787) | 1.476<br>(0.691 to 3.154) | 1.501<br>(0.799 to 2.819) | **1.587**<br>**(1.081 to 2.332)** |
| Understand<br>(weighted n=121,262,741) | 1.262<br>(0.481 to 3.314) | 0.584<br>(0.164 to 2.081) | 1.743<br>(0.776 to 3.914) | 1.183<br>(0.706 to 1.982) | **1.442**<br>**(1.041 to 1.998)** |
| Quality<br>(weighted n=121,499,392) | 0.884<br>(0.390 to 2.004) | 1.069<br>(0.079 to 1.037) | 0.950<br>(0.422 to 2.138) | **1.901**<br>**(1.089 to 3.319)** | 1.352<br>(0.935 to 1.956) |

*Note.* Models were implemented using PROC SURVEYLOGISTIC in SAS using replicate weights and complete case analysis. The first outcome (seeking cancer information) is across the entire population. The latter four models (effort, frustration, understanding, quality) are conditional on seeking cancer information; estimates for these barriers should be interpreted as conditional odds ratios.
**Bold** is significant prior to Holm-Bonferroni within-outcome family adjustment. \* indicates significant at 0.05 level after Holm-Bonferroni within-outcome family adjustment.

Unweighted sensitivity analyses, representing the sample-level rather than population-wide associations, are in **Supplemental Appendix 1**. These analyses indicate that people who are deaf or hard-of-hearing, blind/low vision, or have multiple disabilities have lower odds of seeking cancer-related information. Among those who seek cancer information, there are disparities among people who are blind/low vision (in effort, understanding, and quality), those with mobility disabilities (understanding), and people with chronic pain (in effort and frustration). Those with multiple disabilities had higher adjusted odds of all four information barriers. The differences between weighted and unweighted results illustrate the distinction between population-generalizable effects (weighted) and sample-level associations (unweighted). Weighted models account for survey design and produce estimates representative of U.S. adults with disabilities, whereas unweighted models describe patterns observed in the analytic sample, which may overstate precision for small subgroups. The unweighted results can identify potential disparities for further study, but the weighted results provide the most reliable evidence for population-level inference.

## Discussion

People with disabilities (PWDs) are a growing population who have increased health inequities that could be minimized by accessible health information. HINTS – a yearly, nationally representative survey focused on consumer health information in the U.S. – included options for four different types of disability for the first time in 2024. This novelty allows for analysis on whether people with different types of disabilities experience seeking health information differently. The aim of this study was to determine whether health information seeking experiences, specifically barriers, differed based on type of disability. Our hypothesis was partially supported since deaf individuals had worse health information outcomes, but people who are blind/low vision did not. However, we found that people with multiple disabilities experienced the most disparities when seeking and looking for health information.

The largest group of PWDs in our study were people with multiple disabilities; the most frequent co-occurrence of disability was chronic pain and mobility disability. This group also experienced significant information barriers. People with multiple disabilities had lower odds of seeking cancer-related health information (as were those who were DHH). If information was sought, those with multiple disabilities had higher odds to report feeling frustrated during the search and difficulty understanding the information they found. Other disabilities, such as chronic pain, had higher odds of being concerned about the quality of information that was found. Despite people with multiple disabilities (e.g., those with both chronic pain and mobility disabilities) experiencing information barriers, those with only mobility impairments and chronic pain did not have higher odds of these barriers. This finding emphasizes that although any disability may have a negative effect on a person’s health information seeking experiences, the presence of more than one disability greatly increases the risk of those negative experiences. This pattern is similar to findings of poorer health status^15-16^, health literacy, and socioeconomic position among people with multiple disabilities, and indicates that these people may require more individualized approaches to healthcare and communication.^17^

### Limitations

HINTS 7 is a nationally representative survey but there are still sampling concerns, especially when analyzing responses from people with disabilities. The main concern is that people with “severe” disabilities, such as those in care homes, may not be recruited or eligible for recruitment resulting in a selection bias. Confounding this problem, since this survey is mainly administered through postal services there are issues with accessibility of both survey distribution and the survey itself. Those who use ASL or read Braille face access issues to taking surveys.^18-19^ This issue results in a further selection bias, where those who use alternative modes of communication cannot take the survey unassisted. Consequently, there is less representation of PWDs who are even more likely to have issues seeking health information due to the severity of their disability. Plainly, the samples are not representative of populations that would show the most disparities, contributing to the “no data, no problem” problem.^20^ Still, the inclusion of these disabilities for the first time in HINTS permitted an analysis that suggests widespread information marginalization among PWDs. This finding is supported by the weighted data, providing more conservative estimates (due to weights increasing standard errors and potentially inflating *p*-values), and in the unweighted, sample-specific (sensitivity) analyses. These findings highlight the importance of including disability status when measuring health behavior outcomes in national surveillance studies.

An additional limitation, common with existing data, was he limited number of survey items about disability and health information seeking experiences. Notably, there was no “other” option for disability – there was no data for those with developmental, intellectual, or other disabilities, which are more prevalent than sensory disabilities. Future studies should provide more accessible formats and provide more inclusive (and more precise) measurement of disability.

## Conclusion

We found that PWDs, specifically people who are deaf, have multiple disabilities, or have chronic pain, experience disparities to cancer-related health information seeking. Our findings are consistent with broader literature providing evidence of widespread health information marginalization among PWDs. Discrimination and marginalization of PWDs in healthcare often stems from an intersection of individual factors (e.g., health literacy) and structural health system issues (e.g., lack of accommodations for communication access, lack of accessible information).^10,21-22^ Although decreasing the risk of information marginalization requires improving individual-level factors (e.g., health literacy), intervention strategies must focus on the broader health and public health system to ensure PWDs are included in public health surveillance and programming.

## Data Availability

Data are publicly available from the NIH/National Cancer Institute/Health Information National Trends Study.

---

**Supplemental Table 1.** Unweighted adjusted logistic regression results under Full Information Maximum Likelihood implemented in Mplus.

|  | Deaf | Blind | Mobility | Pain | Multiple |
| --- | --- | --- | --- | --- | --- |
| Seeking cancer information | **0.578**<br>**(0.415 to 0.806)** | **0.582**<br>**(0.419 to 0.810)** | 0.763<br>(0.578 to 1.008) | 0.852<br>(0.708 to 1.026) | **0.820**<br>**(0.704 to 0.955)** |
| Effort | 1.230<br>(0.731 to 2.070) | **1.907**<br>**(1.160 to 3.136)** | 1.374<br>(0.917 to 2.058) | **1.412**<br>**(1.092 to 1.825)** | **1.628**<br>**(1.324 to 2.002)** |
| Frustration | 1.418<br>(0.848 to 2.371) | 1.611<br>(0.980 to 2.647) | 1.242<br>(0.819 to 1.884) | **1.522**<br>**(1.175 to 1.972)** | **1.639**<br>**(1.328 to 2.021)** |
| Understand | 1.246<br>(0.740 to 2.098) | **1.763**<br>**(1.071 to 2.901)** | **1.646**<br>**(1.092 to 2.479)** | 1.118<br>(0.861 to 1.451) | **1.445**<br>**(1.173 to 1.781)** |
| Quality | 0.966<br>(0.582 to 1.603) | **1.781**<br>**(1.038 to 3.057)** | 0.946<br>(0.629 to 1.422) | 1.278<br>(0.983 to 1.660) | **1.363**<br>**(1.102 to 1.686)** |

